# Interstitial Fibrosis and Arrhythmic Mitral Valve Prolapse: Unravelling Sex-Based Differences

**DOI:** 10.1101/2024.01.12.24301217

**Authors:** Lionel Tastet, Shalini Dixit, Thuy Nguyen, Lisa J. Lim, Mohammad Al-Akchar, Dwight Bibby, Farzin Arya, Luca Cristin, Shafkat Anwar, Satoshi Higuchi, Henry Hsia, Yoo Jin Lee, Francesca N. Delling

## Abstract

**Background:** Interstitial fibrosis as quantified by cardiac magnetic resonance (CMR) has been demonstrated in arrhythmic mitral valve prolapse (MVP), a condition with known female predominance. However, prior studies included only MVP cases with significant mitral regurgitation (MR) or mitral annular disjunction (MAD). We sought to evaluate the association between interstitial fibrosis and complex ventricular ectopy (ComVE) in MVPs unselected for MAD or severe MR, and to investigate the contribution of sex to this association.

**Methods:** We performed contrast CMR in consecutive individuals with MVP between 2020 and 2022. Extracellular volume fraction (ECV%), a surrogate marker for interstitial fibrosis, was quantified using T_1_ mapping. Replacement fibrosis was assessed using late gadolinium enhancement (LGE). ComVE, defined as frequent premature ventricular contractions and/or non-sustained/sustained ventricular tachycardia (VT), was detected using ambulatory ECG monitoring.

**Results:** We identified 59 MVP cases without severe MR (49% women, 80% with mild or less MR) and available ECV% measurement. Among these, 23 (39%) had ComVE, including a case of aborted ventricular fibrillation (VF) and one with sudden arrhythmic death, both females. Global ECV% was significantly greater in ComVE versus non-ComVE (31%[27-33] vs 27%[23-30], p=0.002). In MVP-ComVE, higher segmental ECV% was not limited to the inferolateral/inferior LV wall, but was also demonstrated in atypical segments including the anterior/anterolateral wall (p<0.05). The association between ComVE and ECV% was driven by female sex (32%[30-33] vs 28%[26-30], p=0.003 in females; 31%[25-33] vs 26%[23-30], p=0.22 in males). ECV% remained independently associated with an increased risk of ComVE, including VT/VF, after adjustment for cardiovascular risk factors, MAD, and LGE (p<0.01).

**Conclusion:** In MVP without significant MR, interstitial fibrosis by CMR is associated with an increased risk of ComVE, suggesting a primary myopathic process. The stronger association between interstitial fibrosis and ComVE in females may explain why severe arrhythmic complications are more prevalent among women.

GRAPHICAL ABSTRACT:
Expansion of Interstitial Myocardial Fibrosis in Mitral Valve Prolapse with Complex Ventricular Ectopy. Illustrative MVP case with ComVE and greater interstitial fibrosis as demonstrated by T_1_ mapping on CMR (Top). The violin plot shows significantly greater ECV% in MVPs with ComVE compared to non-ComVE (Top). The association between greater interstitial fibrosis (i.e. ECV%) and ComVE was stronger in women than men (Bottom). MR = mitral regurgitation

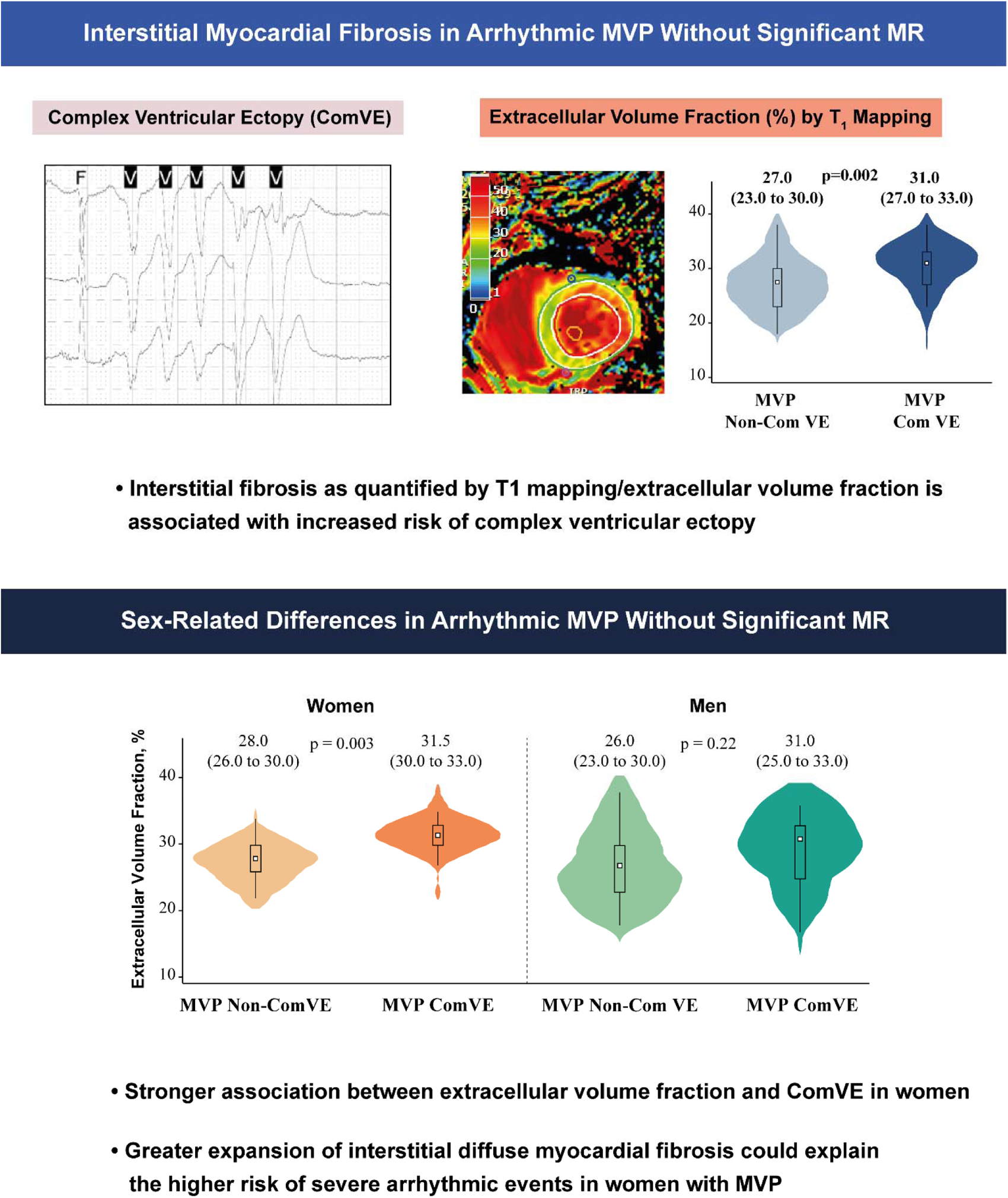

## INTRODUCTION

Mitral valve prolapse (MVP) is a common heritable condition affecting 2% to 3% of individuals worldwide.^1^ MVP without significant mitral regurgitation (MR) is generally considered a benign condition. However, a small but non-negligible subset of MVP subjects (0.14% to 1.8% yearly) are at risk of life-threatening arrhythmic events and sudden cardiac death (SCD).^2, 3^ This “malignant” form of MVP has been commonly associated with female sex, younger age, bileaflet involvement with multi-segmental myxomatous disease, replacement fibrosis in the papillary muscles and adjacent left ventricular (LV) wall, and complex ventricular ectopy (ComVE).^3–5^ ComVE, defined as non-sustained or sustained ventricular tachycardia (VT) and/or frequent pleomorphic PVCs, may be found even in MVP without significant MR and is associated with excess mortality.^5–7^

The role of myocardial fibrosis as a substrate for ComVE and SCD in MVP is supported by seminal histologic and imaging studies.^8–11^ Indeed, MVP-related abnormal valvular-myocardial mechanics related to greater prolapse, mitral annular disjunction (MAD), excessive displacement of papillary muscles, and curling of the basal myocardium may induce myocardial fibrosis.^12^ Replacement fibrosis, typically involving the papillary muscles and LV inferolateral wall, can be detected by late gadolinium enhancement (LGE) on cardiac magnetic resonance (CMR) and relates to arrhythmic MVP.^9, 13–15^ However, ComVE may be present even in the absence of replacement myocardial fibrosis.^10, 16^ In addition to LGE, CMR can detect diffuse or interstitial fibrosis using the extracellular volume fraction (ECV%) calculated from T_1_ mapping.^17, 18^ As shown by our group in a post-mortem investigation^19^, diffuse interstitial myocardial fibrosis could represent an alternative substrate for life-threatening arrhythmia in MVP^20^, regardless of MR severity or replacement fibrosis. Prior studies investigating the role of T_1_ mapping/ECV% in arrhythmic MVP have included mostly cases with severe MR, limiting our understanding about the presence of a primary myopathic process in MVP.^20, 21^ Furthermore, it remains uncertain whether sex-related differences, which may impact not only the mitral valve apparatus, but also myocardial structure and function, could influence the contribution of diffuse myocardial fibrosis to ComVE in MVP.^22^

We therefore hypothesized that among MVPs without significant MR: ***1)*** interstitial fibrosis as quantified by global ECV% on CMR is increased in those with ComVE, suggesting a primary myopathy in MVP; and ***2)*** the relationship between interstitial fibrosis and ComVE is influenced by female sex.

## METHODS

### Study Population

We identified 113 consecutive MVPs recruited as part of the University of California San Francisco (UCSF) MVP Registry from 2020 to 2022 with adequate contrast CMR/T1 mapping and without the following: i) severe MR and/or prior mitral valve (MV) intervention; ii) ischemic or non-ischemic cardiomyopathy (including sarcoidosis); iii) LV ejection fraction <35%; iv) complex congenital heart disease; v) >mild aortic valve disease; or vi) prior defibrillator implantation. Eleven studies were excluded because of the lack of comprehensive ambulatory ECG monitoring to ascertain ventricular arrhythmia. All remaining 59 MVPs had complete demographic/ clinical variables within a calculated Charlson comorbidities index,^23^ and laboratory, ambulatory ECG and CMR data. Ethics Committee of UCSF approved the study and participants gave informed consent. All data produced in the present study are available upon reasonable request to the authors.

### Ascertainment of Ventricular Arrhythmia

Ventricular arrhythmia data were ascertained through 48-hour Holter monitors. ComVE was defined as: ≥5% burden of pleomorphic premature ventricular contractions (PVCs) (isolated and/or in couplets/triplets), presence of sustained or non-sustained ventricular tachycardia (NSVT), or ventricular fibrillation (VF).^7, 16^ Arrhythmic cause of death was ascertained using the National Death Index for SCD cases. PVC burden and origin (i.e., papillary muscles, mitral valve/annulus, LV/RV outflow tract, or other) were also recorded. Pleomorphic ventricular ectopy was defined as PVCs of at least 2 different morphologies. NSVT was defined as > 3 PVCs with a rate >100 bpm lasting <30 seconds.^16^

### Cardiac Magnetic Resonance Imaging

#### Acquisition

Imaging was performed using the UCSF Core using 3.0T scanners Discovery MR750w or SIGNA Premier (GE Healthcare, Milwaukee, WI).^24^ Briefly, standard long- and short-axis cine images covering the entire LV were obtained using a balanced steady-state free precession sequence. Typical parameters were TR/TE 2.8/1.3ms, flip angle 45°. LGE was performed 5-15 min following injection of 0.1 mmol/kg gadobutrol, and images were obtained using a high-resolution breath-hold two-dimensional sequence at three separate levels in the short-axis plane (basal, mid, and apical). An inversion-recovery fast gradient-echo sequence was performed in two phase-encoding directions to differentiate true late enhancement from artefact.^25, 26^ The inversion time was optimized to achieve satisfactory nulling of the myocardium. T_1_ mapping data were acquired using a Modified Look-Locker Inversion-recovery (MOLLI) sequence with built-in motion correction and an acquisition scheme of 3(3)-3(3)-5 both before and following contrast agent administration. Two-dimension (2D) phase-contrast (through-plane), velocity-encoded, imaging approach through the aortic valve was performed to quantify MR.

#### Image Analysis

Image analysis was performed offline using a standardized approach (cvi42 version 5.14.0, Circle Cardiovascular Imaging Inc., Calgary, Canada). LV volume and mass were indexed to body surface area. Papillary muscles were included when measuring LV mass and excluded when measuring volumes.^26^ MVP was defined as superior displacement of one or both mitral leaflets >2 mm beyond the mitral annulus on long axis view, as previously described.^8^ The MVP phenotype was further subdivided as: bileaflet or single-leaflet (anterior or posterior). MR volume was calculated as the difference between LV stroke volume and the forward aortic flow volume. MR fraction was obtained by dividing MR volume by the LV stroke volume. MR severity was assessed using on a qualitative and/or quantitative approach, with visual assessment or 2D phase-contrast imaging when available.

The presence of MAD (inferolateral vs inferior, anterior, or anterolateral) was assessed qualitatively on cine long-axis views as a separation between the left atrial wall at the level of MV junction and the LV free wall.^27^ In addition, LV “curling”, defined as an abnormal or exaggerated basal myocardial systolic motion, was qualitatively and quantitatively assessed as previously described.^12, 14^

On dedicated images, LGE was quantified in a semi-automated manner using a signal intensity threshold of >5 standard deviations above the mean value in a region of normal myocardium. Areas of inversion artefact or signal contamination by epicardial fat or blood pool were manually excluded. LGE quantification was expressed as percentage of LV mass.

T_1_ mapping data were generated from a short-axis cine stack of the LV, and the regions of the endocardium and epicardium on the native T1 maps were manually contoured to avoid partial-volume or cardiac-motion-related blurring of the myocardium-blood boundary, where blood signal may contaminate myocardial data. The LV mid-section was selected due to its greater reproducibility and use in prior MVP-based CMR studies.^21, 28^ A 10% offset was also applied to avoid signal contamination from the blood pool and epicardial fat. The pre-contrast contours were then copied onto the corresponding post-contrast maps with minor adjustments made to avoid partial volume effects. Regions of interest were then drawn in the blood, avoiding myocardium and papillary muscles. ECV fraction was calculated according to the following formula: ECV%= partition coefficient x [1-hematocrit], where partition coefficient = [ΔR1_myocardium_/ΔR1_blood-pool_] and ΔR1 = (1/post-contrast T1-1/native T1).^29^ Hematocrit was measured as per protocol immediately before CMR study. Native myocardial T1 and ECV% were also recorded for all myocardial segments, including the LV inferolateral wall, a location typically subject to abnormal traction by the prolapsing leaflets.^14, 21, 30^

### Statistical Analysis

Continuous variables were expressed as mean ± SD or median (interquartile range [IQR]), and were tested for normality of distribution and homogeneity of variances with the Shapiro-Wilk and Levene tests, respectively. Continuous variables with normal distribution were compared between groups with Student’s t test. Continuous variables non-normally distributed were compared with Wilcoxon-Mann-Whitney test. Categorical variables were presented as frequencies and percentages and were compared with ^2^ test or Fisher’s exact test as appropriate. Univariable and multivariable linear regression analyses were performed to identify the factors significantly associated with diffuse myocardial fibrosis (i.e. ECV%). The multivariable model included all clinically relevant factors and those with a p-value <0.20 in univariable analysis to predict ECV% (i.e. age, inverted lateral T waves, MR severity, LV end-systolic volume index, LGE, and hematocrit). Results were presented as standardized beta coefficient (β) with standard error. A two-tailed p value ≤0.05 was considered significant. Statistical analyses were performed with Stata software version 14.2 (StataCorp, College Station, Texas).

## RESULTS

### Study Population

Demographic, arrhythmic, and CMR characteristics of the study population are presented in **Table 1**. Of the 59 MVPs included in this analysis, a total of 23 (39%) cases (48% women, and 30% men) had ComVE. Among these, one later experienced sudden arrhythmic death, and another had an aborted VF arrest treated with an implantable cardiac defibrillator (ICD). Both were females.

**TABLE 1:** Characteristics of the Study Population.

|  | All Patients<br>(N = 59) | MVP ComVE<br>(n = 23; 39%) | MVP Non-ComVE<br>(n = 36; 61%) | p Value |
| --- | --- | --- | --- | --- |
| ***Demographic data*** |  |  |  |  |
| Age, years | 54 ± 15 | 57 ± 16 | 53 ± 14 | 0.29 |
| Female sex, n (%) | 29 (49) | 14 (60) | 15 (41) | 0.15 |
| Body surface area, m<sup>2</sup> | 1.82 ± 0.19 | 1.82 ± 0.20 | 1.82 ± 0.18 | 0.94 |
| Symptoms, n (%) | 16 (27) | 8 (35) | 8 (22) | 0.29 |
| Dyspnea, n (%) | 2 (3) | 2 (8) | 0 (0) | 0.14 |
| Chest pain, n (%) | 3 (5) | 2 (8) | 1 (2) | 0.31 |
| Palpitations, n (%) | 9 (15) | 3 (13) | 6 (16) | 1.00 |
| Syncope, n (%) | 4 (6) | 2 (8) | 2 (5) | 0.63 |
| Cardiac arrest/SCD, n (%) | 1 (1) | 1 (4) | 0 (0) | - |
| Family history of SCD, n (%) | 9 (15) | 5 (22) | 4 (11) | 0.28 |
| ***ECG/Holter data*** |  |  |  |  |
| Pleomorphic ventricular ectopy, n (%) | 33 (58) | 15 (71) | 18 (51) | 0.14 |
| Non-sustained VT, n (%) | 19 (34) | 19 (90) | 0 (0) | - |
| PVCs burden, (%) | 1.6 ± 3.6 | 3.4 ± 5.1 | 0.3 ± 0.5 | **0.001** |
| PVCs burden ≥5%, n (%) | 5 (12) | 5 (33) | 0 (0) | - |
| PVCs by origin |  |  |  | **0.02** |
| Posterior papillary muscles, n (%) | 11 (25) | 7 (47) | 4 (13) |  |
| Anterior papillary muscles, n (%) | 8 (18) | 2 (13) | 6 (21) |  |
| Anterior/posterior MV, n (%) | 2 (5) | 2 (13) | 0 (0) |  |
| LV/RV outflow tract, n (%) | 7 (16) | 1 (7) | 6 (21) |  |
| Other, n (%) | 16 (36) | 3 (20) | 13 (45) |  |
| Inverted T waves, n (%) | 20 (35) | 9 (39) | 11 (33) | 0.65 |
| Inverted inferolateral T waves, n (%) | 3 (5) | 1 (4) | 2 (6) | 1.00 |
| Inverted inferior T waves, n (%) | 23 (40) | 11 (47) | 12 (35) | 0.34 |
| Inverted lateral T waves, n (%) | 1 (2) | 0 (0) | 1 (3) | 1.00 |
| ***CMR data*** |  |  |  |  |
| MVP subtype |  |  |  | 0.24 |
| Bileaflet, n (%) | 38 (64) | 18 (78) | 20 (55) |  |
| Anterior, n (%) | 8 (13) | 2 (8) | 6 (16) |  |
| Posterior, n (%) | 13 (22) | 3 (13) | 10 (27) |  |
| No or mild MR, n (%) | 47 (80) | 19 (83) | 28 (78) | 0.75 |
| Mitral annular disjunction |  |  |  |  |
| Any site, n (%) | 39 (66) | 16 (70) | 23 (64) | 0.65 |
| Inferolateral, n (%) | 30 (51) | 11 (48) | 19 (54) | 0.63 |
| LV end-diastolic volume index, ml/m<sup>2</sup> | 83 ± 18 | 82 ± 16 | 83 ± 20 | 0.80 |
| LV end-systolic volume index, ml/m<sup>2</sup> | 32 ± 10 | 33 ± 10 | 31 ± 10 | 0.47 |
| LV ejection fraction, % | 61 ± 6 | 59 ± 7 | 62 ± 5 | 0.05 |
| LV mass index, g/m<sup>2</sup> | 45 ± 14 | 48 ± 10 | 44 ± 16 | 0.31 |
| LV systolic curling, n (%) | 36 (66) | 16 (76) | 20 (60) | 0.23 |
| LV systolic curling, mm | 3.8 ± 1.7 | 4.0 ± 2.0 | 3.5 ± 1.5 | 0.40 |
| LGE present, n (%) | 19 (30) | 8 (35) | 11 (30) | 0.73 |
| Inferolateral LV wall, n (%) | 9 (15) | 3 (13) | 6 (16) | 1.00 |
| Papillary muscles, n (%) | 6 (10) | 3 (13) | 3 (8) | 0.66 |
| T<sub>1</sub> mapping mid LV |  |  |  |  |
| Native T<sub>1</sub>, ms | 1261 ± 49 | 1284 ± 10 | 1247 ± 43 | **0.004** |
| Post contrast T<sub>1</sub>, ms | 570 ± 64 | 554 ± 72 | 580 ± 57 | 0.13 |
| Hematocrit, % | 42.5 ± 3.6 | 42.5 ± 3.1 | 42.6 ± 3.9 | 0.94 |
| Extracellular volume fraction, % |  |  |  |  |
| Mean, % | 28.4 ± 4.8 | 30.3 ± 4.7 | 27.2 ± 4.6 | **0.01** |
| Median, % | 29.0 (24.0 - 32.0) | 31.0 (27.0 - 33.0) | 27.5 (23.0 - 30.0) | **0.006** |
| Maximum ECV inferolateral, n (%) | 33 (56) | 16 (69) | 17 (48) | 0.11 |

Values are mean ± SD or median (25<sup>th</sup> – 75<sup>th</sup> percentiles).
CMR = cardiac magnetic resonance; ECG = electrocardiogram; ECV = extracellular volume; LGE = late gadolinium enhancement; LV = left ventricular; MR = mitral regurgitation; MV = mitral valve; MVP = mitral valve prolapse; PVC = premature ventricular contraction; VT = ventricular tachycardia; SCD = sudden cardiac death.

PVC burden was significantly higher in the ComVE group, and by definition all cases in this group had PVC burden ≥5% (**Table 1**). PVCs were pleomorphic in all cases with PVCs burden ≥5%, and originated from typical MVP sites such as the papillary muscles and MV annulus^31^ (**Table 1**). The proportion of T-wave inversion, including inverted inferolateral T-waves, was comparable between groups (**Table 1**).

CMR characteristics, including MR severity (mild or less versus moderate), LV volumes, presence of MAD (inferolateral: 48% vs 54%, p=0.63) and LGE in the LV inferolateral segment (13% vs 16%, p=1.00) and papillary muscles (13% vs 8%, p=0.66), were comparable between groups (**Table 1**). However, the ComVE group had lower LV ejection fraction (59±7% vs 62±5%, p=0.05) and a trend toward more bileaflet involvement (78% vs 55%, p=0.07) compared to the non-ComVE group.

### Ventricular Arrhythmia, Diffuse Interstitial Fibrosis and Sex-Based Differences

Native T1 was greater in the ComVE than non-ComVE group (1284±10 vs 1247±43 ms, p=0.004), while post-contrast T1 was slightly lower in the ComVE group (554±72 vs 580±57 ms, p=0.13). Mean hematocrit value was similar between groups (42.5±3.1 vs 42.6±3.9 ms, p=0.94).

Median global ECV% was significantly greater in MVP with ComVE compared to non-ComVE (31.0% [27.0-33.0] vs 27.5% [23.0-30.0], p=0.006) (**Figure 2**). Moreover, exclusion of MVPs with LGE (n=19) provided similar results (31.0% [27.0-33.0] vs 27.0% [23.0-30.0], p=0.01). Regardless of ComVE, women demonstrated greater ECV% compared to men (30.0% [27.0-32.0] vs 27.0% [23.0-32.0]), although the comparison was not statistically significant (p=0.10). Importantly, women with ComVE had significantly greater ECV% than those without ComVE (31.5% [30.0-33.0] vs 28.0% [26.0-30.0], p=0.003) (**Figure 2**). In men, there was no significant difference in ECV% between ComVE and non-ComVE (31.0% [25.0-33.0] vs 27.0% [23.0-30.0], p=0.32) (**Figure 2**).

**FIGURE 1:**
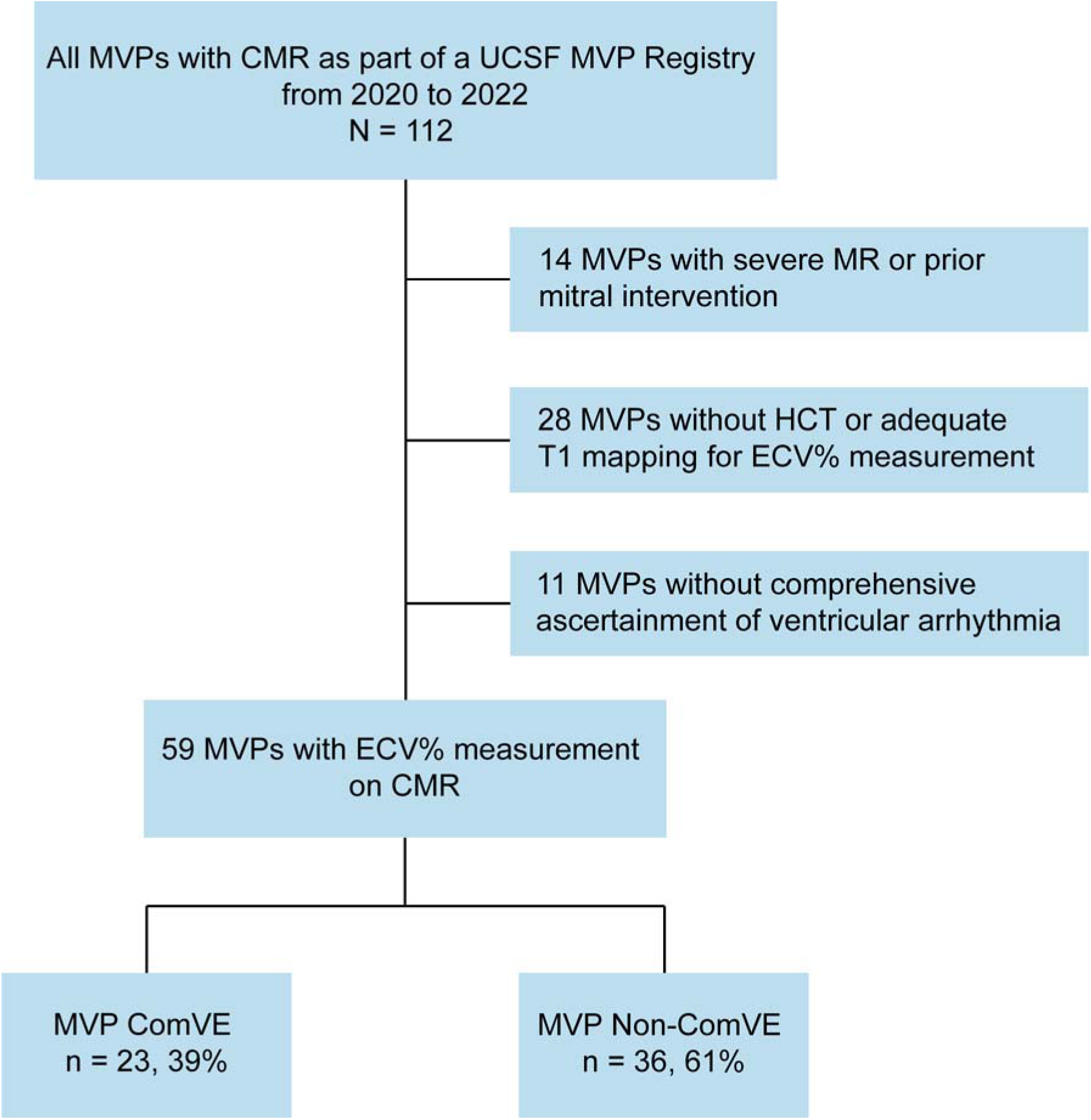
Study Flow Chart. CMR = cardiac magnetic resonance; ComVE = complex ventricular ectopy; ECV = extracellular volume; ECV% = ECV fraction; HCT = hematocrit; MR = mitral regurgitation; MVP = mitral valve prolapse; UCSF = University of California San Francisco.

**FIGURE 2:**
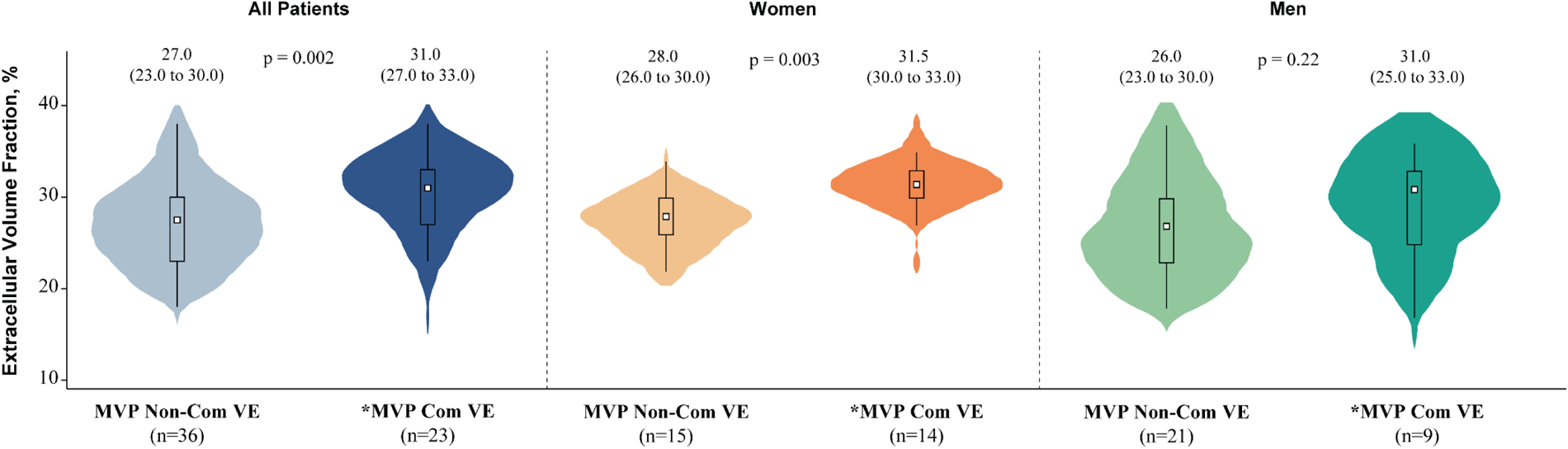
Distribution of Extracellular Volume Fraction According to Complex Ventricular Ectopy. Comparison of ECV% according to ComVE in the whole cohort, subset of women, and men. The violin plot shows the median value (white square), the interquartile range (IQR), the error bars, and the kernel density plot. The numbers on top of the dot plot indicate the median with IQR between brackets. The numbers between brackets below the graph indicate the number of patients for each subgroup. Abbreviation as in Figure 1.

Figure 3 shows the distribution of ECV% across the mid LV segments between ComVE and non-ComVE. Median ECV% was significantly greater in ComVE versus non-ComVE across all segments (all p≤0.05), except for the anteroseptal segment (Figure 3A). Furthermore, in MVPs with ComVE, median ECV% was significantly increased in the inferolateral segment compared to the other segments (all p≤0.05), with the exception of the inferior segment (Figure 3A). MVPs without ComVE had a different distribution of ECV%, with no increase in median ECV% in the inferolateral segment compared to other myocardial segments (Figure 3A). Similarly, women with ComVE had significantly greater ECV% across all segments compared to those without ComVE (all p≤0.05), except for the anteroseptal and anterior segments (Figure 3B). Moreover, median ECV% in the inferolateral segment was markedly increased compared to other myocardial segments in women with ComVE, but not in those without ComVE (Figure 3B). In the subset of men, there were no significant differences in median ECV% across all myocardial segments between ComVE versus non-ComVE (Figure 3C). Moreover, in the ComVE group, median ECV% in the inferolateral segment was significantly increased only when compared to the anterolateral segment. In the non-ComVE group, median ECV% in the inferolateral segment was not significantly increased compared to the other myocardial segments (Figure 3C).

**FIGURE 3:**
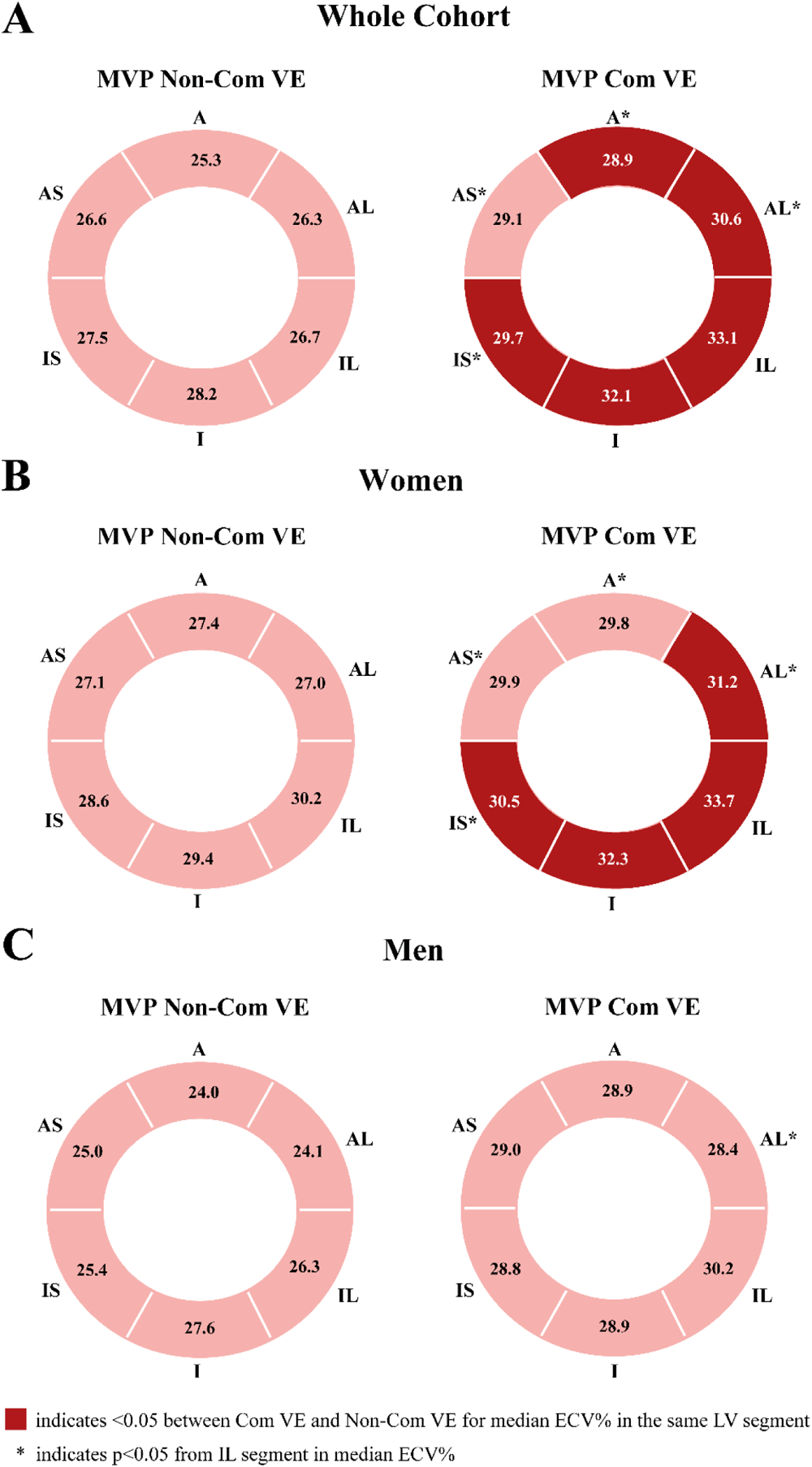
Distribution of Extracellular Volume Fraction in Myocardial Segments According to Complex Ventricular Ectopy. Comparison of ECV% in the mid LV myocardial segments according to ComVE in the whole cohort (A), subset of women (B), and men (C). A = anterior; AL = anterolateral; AS = anteroseptal; I = inferior; IL = inferolateral; IS = inferoseptal; other abbreviation as in Figure 1.

**Table 2** shows ECG and CMR characteristics according to ComVE and sex. Overall, there were no significant differences between groups, with the exception of higher global native T_1_ (1302±49 vs 1249±50 ms, p=0.008) and inferolateral native T_1_ (1335±73 vs 1263±78 ms, p=0.01) in women with ComVE compared to those without ComVE. In men, global native T_1_ (1255±55 vs 1245±39 ms, p=0.52) and inferolateral native T_1_ (1292±38 vs 1277±60 ms, p=0.48) were similar between ComVE and non-ComVE. MVPs with ComVE were more likely to have bileaflet involvement and lower LVEF in both women and men (**Table 2**). The proportion of MVPs with MAD and/or LGE did not differ between groups among women and men (**Table 2**).

**TABLE 2:** ECG and CMR Characteristics According to Sex and Arrhythmia.

|  | Women |  |  | Men |  |  |
| --- | --- | --- | --- | --- | --- | --- |
|  | MVP ComVE<br>(n = 14; 48%) | MVP Non-ComVE<br>(n = 15; 52%) | p Value | MVP ComVE<br>(n = 9; 30%) | MVP Non-ComVE<br>(n = 21; 70%) | p Value |
| ***ECG/Holter data*** |  |  |  |  |  |  |
| Pleomorphic ventricular ectopy, n (%) | 7 (54) | 7 (50) | 0.84 | 8 (89) | 11 (52) | **0.01** |
| Non-sustained VT, n (%) | 12 (92) | 0 (0) | - | 7 (87) | 0 (0) | - |
| PVCs burden, (%) | 2.7 ± 4.5 | 0.2 ± 0.3 | 0.10 | 4.5 ± 6.1 | 0.3 ± 0.5 | **0.004** |
| PVCs burden ≥5%, n (%) | 2 (29) | 0 (0) | - | 3 (38) | 0 (0) | - |
| PVCs by origin |  |  | 0.73 |  |  | 0.07 |
| Posterior papillary muscles, n (%) | 3 (43) | 2 (20) |  | 4 (50) | 2 (10) |  |
| Anterior papillary muscles, n (%) | 1 (14) | 2 (20) |  | 1 (12) | 4 (21) |  |
| Anterior/posterior MV, n (%) | 1 (14) | 0 (0) |  | 1 (12) | 0 (0) |  |
| LV/RV outflow tract, n (%) | 0 (0) | 1 (10) |  | 1 (13) | 5 (26) |  |
| Other, n (%) | 2 (29) | 5 (50) |  | 1 (13) | 8 (42) |  |
| Inverted T waves, n (%) | 6 (42) | 5 (35) | 0.69 | 3 (33) | 6 (31) | 0.92 |
| Inverted inferolateral T waves, n (%) | 1 (7) | 1 (7) | 1.00 | 0 (0) | 1 (5) | 0.48 |
| Inverted inferior T waves, n (%) | 7 (50) | 6 (42) | 0.70 | 4 (44) | 6 (30) | 0.67 |
| Inverted lateral T waves, n (%) | 0 (0) | 0 (0) | - | 1 (5) | 0 (0) | 0.48 |
| ***CMR data*** |  |  |  |  |  |  |
| MVP subtype |  |  | 0.28 |  |  | 0.56 |
| Bileaflet, n (%) | 10 (71) | 6 (40) |  | 8 (88) | 14 (66) |  |
| Anterior, n (%) | 2 (14) | 4 (26) |  | 1 (11) | 6 (28) |  |
| Posterior, n (%) | 2 (14) | 5 (33) |  | 0 (0) | 1 (4) |  |
| No or mild MR, n (%) | 12 (86) | 13 (87) | 1.00 | 7 (78) | 15 (71) | 1.00 |
| Mitral annular disjunction |  |  |  |  |  |  |
| Any site, n (%) | 9 (64) | 11 (73) | 0.70 | 7 (77) | 12 (57) | 0.42 |
| Inferolateral, n (%) | 7 (50) | 8 (57) | 0.70 | 4 (44) | 11 (52) | 0.69 |
| LV end-diastolic volume index, ml/m<sup>2</sup> | 78 ± 13 | 79 ± 18 | 0.90 | 86 ± 19 | 85 ± 20 | 0.90 |
| LV end-systolic volume index, ml/m<sup>2</sup> | 30 ± 6 | 28 ± 8 | 0.41 | 36 ± 12 | 33 ± 10 | 0.43 |
| LV ejection fraction, % | 60 ± 6 | 64 ± 4 | 0.08 | 57 ± 9 | 61 ± 6 | 0.14 |
| LV mass index, g/m<sup>2</sup> | 44 ± 8 | 40 ± 11 | 0.25 | 53 ± 11 | 46 ± 18 | 0.34 |
| LV systolic curling, n (%) | 11 (84) | 8 (57) | 0.20 | 5 (62) | 12 (63) | 1.00 |
| LV systolic curling, mm | 4.3 ± 2.2 | 3.4 ± 1.5 | 0.35 | 3.5 ± 1.6 | 3.6 ± 1.6 | 0.89 |
| LGE present, n (%) | 3 (21) | 4 (26) | 1.00 | 5 (55) | 7 (33) | 0.41 |
| Inferolateral LV wall, n (%) | 1 (7) | 2 (13) | 1.00 | 2 (22) | 4 (19) | 1.00 |
| Papillary muscles, n (%) | 1 (7) | 2 (13) | 1.00 | 2 (22) | 1 (4) | 0.20 |
| T<sub>1</sub> mapping mid LV |  |  |  |  |  |  |
| Native T<sub>1</sub>, ms | 1302 ± 49 | 1249 ± 50 | **0.008** | 1255 ± 35 | 1245 ± 39 | 0.52 |
| Post contrast T<sub>1</sub>, ms | 549 ± 84 | 571 ± 67 | 0.43 | 562 ± 53 | 586 ± 50 | 0.24 |
| Inferolateral native T<sub>1</sub>, ms | 1335 ± 73 | 1263 ± 78 | **0.01** | 1292 ± 38 | 1277 ± 60 | 0.48 |
| Hematocrit, % | 42.0 ± 3.5 | 41.0 ± 2.7 | 0.39 | 43.2 ± 2.3 | 43.7 ± 4.2 | 0.77 |

Values are mean ± SD or median (25<sup>th</sup> – 75<sup>th</sup> percentiles).
Abbreviations as in **Table 1**.

### Predictors of Complex Ventricular Ectopy

In an exploratory analysis, we assessed the association of ECV% with the risk of ComVE after adjusting for the Charlson comorbidities index, presence of LGE, bileaflet MVP, inferolateral MAD, and LVEF. In univariable analysis, ComVE risk was only predicted by ECV% (OR: 1.18, 95% CI 1.04 - 1.35; p=0.01). Of note, when stratified based on sex, ECV% was significantly associated with the risk of ComVE in women (OR: 1.44, 95% CI 1.07 - 1.94; p=0.01), but not in men (OR: 1.08, 95% CI 0.93 - 1.26; p=0.31). After multiple multivariable models, ECV% remained independently associated with an increased risk of ComVE in all multivariable models (Figure 4).

**FIGURE 4:**
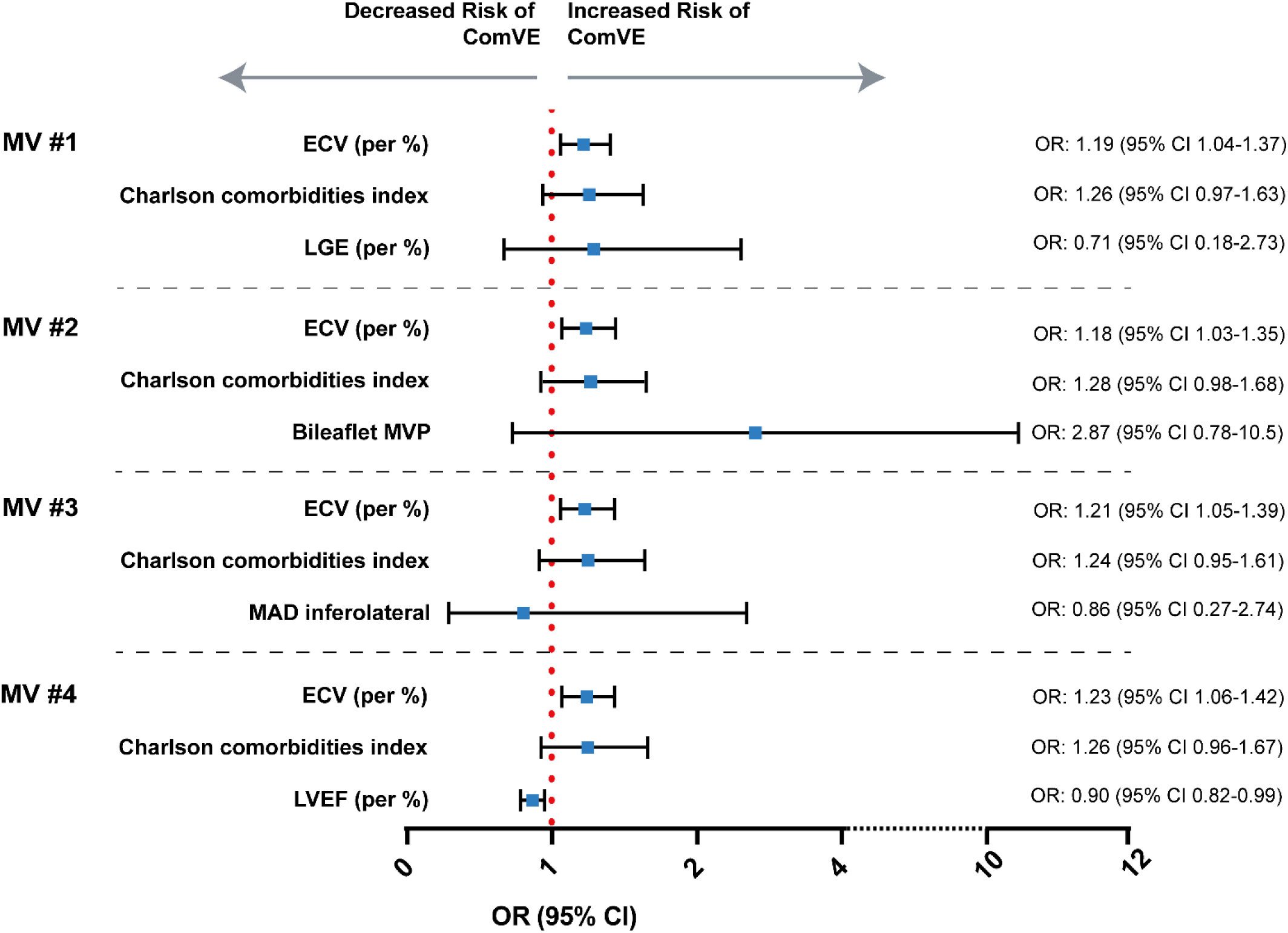
Factors Associated with Risk of Complex Ventricular Ectopy. ECV% remained associated with increased risk of ComVE in multiple multivariable models including several risk factors. CI = confidence interval; LGE = late gadolinium enhancement; LVEF = left ventricular ejection fraction; MAD = mitral annular disjunction; MV = multivariable; OR = odd ratio; other abbreviation as in Figure 1.

## DISCUSSION

In this cross-sectional analysis of consecutive and unselected MVPs with predominantly no or mild MR undergoing CMR, we investigated the relationship of interstitial myocardial fibrosis with ComVE, including cases with VT, VF arrest and SCD. The main findings were as follows: ***1)*** Diffuse interstitial fibrosis as quantified by global ECV%/T_1_ mapping was significantly greater in MVPs with ComVE, regardless of presence of LGE; ***2)*** In the arrhythmic MVP group, higher segmental ECV% was not limited to the inferolateral/inferior LV wall, but was also demonstrated in segments not typically subject to myocardial traction, suggesting a diffuse myopathic process in MVP beyond abnormal valvular-myocardial mechanics; ***3)*** The association between interstitial fibrosis and ComVE was more prominent in women compared to men with MVP (**Graphical Abstract**).

### Myocardial Fibrosis and Arrhythmic MVP

Seminal studies support the role of myocardial fibrosis, resulting from increased mechanical traction of prolapsing valve, as a substrate for ComVE and SCD in MVP.^5, 8, 9, 11^ Basso et al.^9^ previously reported that replacement fibrosis is a common feature of young SCD victims (≤40 years of age) with arrhythmic MVP. However, replacement fibrosis is not consistently found in SCD victims with MVP,^10^ nor in those with ComVE alone (prevalence of LGE 36-45%).^20, 32^ These findings suggest that other myocardial substrates are contributing to arrhythmic outcomes in MVP.

Non-invasive measurement of diffuse interstitial fibrosis can be achieved by measuring ECV% by T_1_ mapping on CMR, a technique that has been validated against histology.^33, 34^ Prior studies have reported abnormal myocardial T_1_ and ECV%, suggestive of greater diffuse interstitial fibrosis in arrhythmic MVP.^21, 28, 35^ However, such investigations have included predominantly MVP cases with severe MR or MAD, or have linked abnormal ECV% to a few cases of sudden cardiac arrest rather than the wider spectrum of ComVE.^20, 21^ ComVE is frequently detected (>80%) in patients with MVP before lethal events and has been linked to mortality excess.^5, 6^ Therefore, the identification of imaging parameters able to predict ComVE are essential for detection of intermediate categories of risk and overall improved arrhythmic risk stratification. Selection of specific categories of MVP in prior studies (severe MR or MVP with MAD and abnormal valve-myocardial mechanics) have limited our understanding of the distribution and pattern of interstitial fibrosis in MVP beyond a simple response to chronic volume load or a focal fibrotic response to myocardial traction. Another study has assessed ECV% in mild or moderate MR, but only in 12 MVP patients and without investigating segmental contributions to global ECV%.^36^ In the present study, we demonstrate for the first time in a larger sample of CMR studies unselected for severe MR or MAD, that ComVE in MVP is strongly linked to greater ECV%, even in the subset without LGE or abnormal valvular-myocardial mechanics. Our imaging findings are in line with our prior post-mortem data demonstrating that multisite interstitial fibrosis was more prevalent than replacement fibrosis (100% vs 30%) on autopsy in consecutive and unselected MVPs with sudden arrhythmic death adjudication.^10^ Therefore, diffuse interstitial fibrosis may represent an alternative substrate for ComVE and SCD in MVP with mild MR, even in the absence of LGE. As such, MVP patients undergoing CMR for assessment of more traditional parameters of risk such as MAD or LGE, may greatly benefit from routine T_1_ mapping/ECV% calculation as part of their clinical care.

To date, the exact mechanisms for ventricular arrhythmia and SCD in MVP without severe MR are not well understood. It has been hypothesized that the arrhythmogenic risk associated with bileaflet MVP with mild MR could be attributed to stretching forces exerted on the papillary muscles and adjacent LV wall by the prolapsing leaflets.^11, 12^ These mechanical stresses could trigger myocardial inflammation and contribute to fibrosis as a substrate for ventricular arrhythmia.^36^ Moreover, MAD, which is more common in bileaflet MVP,^37^ can augment such abnormal valve-myocardial mechanics. In the present study, we confirmed the regionalized expansion of myocardial fibrosis in arrhythmic MVP. Among MVP ComVE, the inferolateral segment had the highest ECV% within the LV mid-section. Further, the absolute ECV% in the LV inferolateral segment was significantly greater in MVP with ComVE compared to non-ComVE. However, among MVP patients with mild MR, we also highlight a global, multi-segment pattern of interstitial fibrosis which may underline a diffuse myopathic process initially proposed in genetic studies highlighting mutations in cardiomyopathy genes (*FLNC, LMNA, ALPK3*A).^38, 39^ A primary myopathy may represent the “substrate” for ventricular arrhythmia in the presence of bileaflet MVP, MAD and myocardial stretch (the “trigger”). Alternatively, such diffuse myopathy may act alone, when bileaflet MVP and associated imaging arrhythmic features are absent. Indeed, in our study sample only 50% of MVP patients with mild MR had the traditional phenotype of bileaflet MVP with MAD and curling. Longitudinal studies are needed to determine the prognostic role of CMR-based diffuse interstitial myocardial fibrosis in arrhythmic MVP either as an imaging surrogate of a primary myopathy or, when regionalized to the inferolateral LV, as a precursor of replacement fibrosis.

### Sex Differences, Expansion of Myocardial Fibrosis, and Risk of Arrhythmias

There are conflicting findings regarding the potential effect of sex on MVP arrhythmogenicity. Prior reports of MVP-related SCD/SCA cases have showed a predominance of young women.^5, 9^

In our study, there was a predominance of women with ComVE (60%), which is consistent with prior observations.^40^ Importantly, we observed a stronger association between global ECV% and ComVE in women compared to men. In addition, women with MVP and ComVE had significantly greater ECV% in the LV inferolateral segment but also in other segments typically not subject to myocardial traction. Of note, both women and men had similar distributions of replacement fibrosis as quantified by LGE. To our knowledge, this is the first study to investigate and report sex-related differences in the expansion of myocardial fibrosis as measured by CMR in MVP.

Several factors may contribute to sex-specific mechanisms of interstitial fibrosis in arrhythmic MVP including neurohormonal differences, cardiac structural and functional variations, and genetic predisposition.^41, 42^ Sex hormones, particularly estrogen, are well-known for their impact on the myocardium.^43^ In this setting, hormonal fluctuations may contribute to ventricular fibrosis and arrhythmias.^41, 44^ Greater global/diffuse interstitial fibrosis in women with MVP, mild MR, and ComVE may suggest a primary/genetic diffuse myopathy with variable, sex-related penetrance similar to that observed for valvular features, i.e. bileaflet MVP and MAD, which are more prevalent in women.^45^ Indeed, in addition to greater global ECV%, women with arrhythmic MVP in our study exhibited greater “regionalized” interstitial fibrosis, specifically higher ECV% in the mid inferolateral LV, which was not observed in men. Altogether, these findings support the role of sex-specific factors in the development of myocardial fibrosis and subsequent risk of ventricular arrhythmia in MVP. However, further studies are needed to provide a clearer understanding of the effect of sex in arrhythmic MVP.

### Clinical Implications

The identification of those at highest risk for ventricular arrhythmia and SCD within the larger population of benign MVP cases remains a major and unmet challenge.^1^ Current guidelines focus on management of significant MR, with less emphasis on routine monitoring of ComVE in MVP without MR.^46, 47^ In the absence of LGE, arrhythmic risk stratification in MVP is even less clear to the practicing cardiologist. In our study, we demonstrate greater interstitial fibrosis in arrhythmic MVP, regardless of MR or LGE, and more so in women. Hence, ECV% quantified by T_1_ mapping represents an alternative surrogate marker for myocardial fibrosis that is expected to improve risk stratification of “malignant” MVP. Individuals with MVP and abnormal ECV%, particularly women without significant MR and/or LGE, may benefit from closer clinical and imaging monitoring. Furthermore, our findings support the need for better understanding of the underlying sex-specific mechanisms involved in the pathogenesis of arrhythmic MVP.

### Limitations

The retrospective design and small sample size of the present study require additional studies to confirm our findings. Nonetheless, this study is the first to our knowledge to characterize the association between interstitial fibrosis and ComVE, in a large series of MVPs undergoing CMR studies unselected for severe MR or ventricular arrhythmia. Importantly, our findings suggest for the first-time sex differences in patterns of fibrosis in MVP which may explain the higher prevalence of arrhythmic complications in women. Our findings need to be validated in further experimental and longitudinal observational studies.

## CONCLUSION

In the first large series of consecutive MVP patients with mild MR, ComVE is associated with greater interstitial fibrosis as quantified by ECV% using CMR/T1 mapping, even in the absence of LGE. Measurement of ECV% could help enhance arrhythmic risk stratification particularly in female arrhythmic MVP patients, who exhibit greater interstitial fibrosis compared to men. Further studies are needed to determine if sex-specific mechanisms are involved in the pathogenesis of arrhythmic MVP.

## Data Availability

All data produced in the present study are available upon reasonable request to the authors.

## ABBREVIATIONS

MVP: mitral valve prolapse
MR: mitral regurgitation
SCD: sudden cardiac death
LV: left ventricular/ventricle
ComVE: complex ventricular ectopy
LGE: late gadolinium enhancement
CMR: cardiac magnetic resonance
ECV: extracellular volume fraction

## Sources of Funding

This work was supported by the National Institute of Health NHLBI R01HL153447.

## Disclosures

Nothing to disclose.

## Notes

### Competing Interest Statement

The authors have declared no competing interest.

### Author Declarations

Ethics Committee of UCSF approved the study and participants gave informed consent.

## REFERENCES

1. Delling FN, Noseworthy PA, Adams DH, Basso C, Borger M, Bouatia-Naji N, et al. Research opportunities in the treatment of mitral valve prolapse: Jacc expert panel. J Am Coll Cardiol. 2022;80:2331–2347

2. Narayanan K, Uy-Evanado A, Teodorescu C, Reinier K, Nichols GA, Gunson K, et al. Mitral valve prolapse and sudden cardiac arrest in the community. Heart Rhythm. 2016;13:498–503

3. Nalliah CJ, Mahajan R, Elliott AD, Haqqani H, Lau DH, Vohra JK, et al. Mitral valve prolapse and sudden cardiac death: A systematic review and meta-analysis. Heart (British Cardiac Society). 2019;105:144–151

4. Corrado D, Basso C, Nava A, Rossi L, Thiene G. Sudden death in young people with apparently isolated mitral valve prolapse [comment] [see comments]. G Ital Cardiol. 1997;27:1097–1105

5. Sriram CS, Syed FF, Ferguson ME, Johnson JN, Enriquez-Sarano M, Cetta F, et al. Malignant bileaflet mitral valve prolapse syndrome in patients with otherwise idiopathic out-of-hospital cardiac arrest. J Am Coll Cardiol. 2013;62:222–230

6. Essayagh B, Sabbag A, Antoine C, Benfari G, Yang LT, Maalouf J, et al. Presentation and outcome of arrhythmic mitral valve prolapse. J Am Coll Cardiol. 2020;76:637–649

7. Sabbag A, Essayagh B, Barrera JDR, Basso C, Berni A, Cosyns B, et al. Ehra expert consensus statement on arrhythmic mitral valve prolapse and mitral annular disjunction complex in collaboration with the esc council on valvular heart disease and the european association of cardiovascular imaging endorsed cby the heart rhythm society, by the asia pacific heart rhythm society, and by the latin american heart rhythm society. Europace. 2022;24:1981–2003

8. Han Y, Peters DC, Salton CJ, Bzymek D, Nezafat R, Goddu B, et al. Cardiovascular magnetic resonance characterization of mitral valve prolapse. JACC. Cardiovascular imaging. 2008;1:294–303

9. Basso C, Perazzolo MM, Rizzo S, De Lazzari M, Giorgi B, Cipriani A, et al. Arrhythmic mitral valve prolapse and sudden cardiac death. Circulation. 2015;132:556–566

10. Delling FN, Aung S, Vittinghoff E, Dave S, Lim LJ, Olgin JE, et al. Antemortem and post-mortem characteristics of lethal mitral valve prolapse among all countywide sudden deaths. JACC Clin Electrophysiol. 2021;7:1025–1034

11. Morningstar JE, Gensemer C, Moore R, Fulmer D, Beck TC, Wang C, et al. Mitral valve prolapse induces regionalized myocardial fibrosis. J Am Heart Assoc. 2021;10:e022332

12. Nagata Y, Bertrand PB, Baliyan V, Kochav J, Kagan RD, Ujka K, et al. Abnormal mechanics relate to myocardial fibrosis and ventricular arrhythmias in patients with mitral valve prolapse. Circulation. Cardiovascular imaging. 2023;16:e014963

13. Han Y, Peters DC, Kissinger KV, Goddu B, Yeon SB, Manning WJ, et al. Evaluation of papillary muscle function using cardiovascular magnetic resonance imaging in mitral valve prolapse. Am J Cardiol. 2010;106:243–248

14. Perazzolo Marra M, Basso C, De Lazzari M, Rizzo S, Cipriani A, Giorgi B, et al. Morphofunctional abnormalities of mitral annulus and arrhythmic mitral valve prolapse. Circulation. Cardiovascular imaging. 2016;9:e005030

15. Kitkungvan D, Nabi F, Kim RJ, Bonow RO, Khan MA, Xu J, et al. Myocardial fibrosis in patients with primary mitral regurgitation with and without prolapse. J Am Coll Cardiol. 2018;72:823–834

16. Tison GH, Abreau S, Barrios J, Lim LJ, Yang M, Crudo V, et al. Identifying mitral valve prolapse at risk for arrhythmias and fibrosis from electrocardiograms using deep learning. JACC: Advances. 2023;2:100446

17. Taylor AJ, Salerno M, Dharmakumar R, Jerosch-Herold M. T1 mapping: Basic techniques and clinical applications. JACC. Cardiovascular imaging. 2016;9:67–81

18. Everett RJ, Treibel TA, Fukui M, Lee H, Rigolli M, Singh A, et al. Extracellular myocardial volume in patients with aortic stenosis. J Am Coll Cardiol. 2020;75:304–316

19. Tseng ZH, Olgin JE, Vittinghoff E, Ursell PC, Kim AS, Sporer K, et al. Prospective countywide surveillance and autopsy characterization of sudden cardiac death: Post scd study. Circulation. 2018;137:2689–2700

20. Bui AH, Roujol S, Foppa M, Kissinger KV, Goddu B, Hauser TH, et al. Diffuse myocardial fibrosis in patients with mitral valve prolapse and ventricular arrhythmia. Heart (British Cardiac Society). 2017;103:204–209

21. Pavon AG, Arangalage D, Pascale P, Hugelshofer S, Rutz T, Porretta AP, et al. Myocardial extracellular volume by t1 mapping: A new marker of arrhythmia in mitral valve prolapse. J Cardiovasc Magn Reson. 2021;23:102

22. DesJardin JT, Chikwe J, Hahn RT, Hung JW, Delling FN. Sex differences and similarities in valvular heart disease. Circ Res. 2022;130:455–473

23. Charlson M, Pompei P, Ales K, MacKenzie C. A new method of classifying prognostic comorbidity in longitudinal studies: Development and validation. J Chronic Dis. 1987;40:373–383

24. Marano PJ, Lim LJ, Sanchez JM, Alvi R, Nah G, Badhwar N, et al. Long-term outcomes of ablation for ventricular arrhythmias in mitral valve prolapse. J Interv Card Electrophysiol. 2021;61:145–154

25. Biglands JD, Radjenovic A, Ridgway JP. Cardiovascular magnetic resonance physics for clinicians: Part ii. J Cardiovasc Magn Reson. 2012;14:66

26. Schulz-Menger J, Bluemke DA, Bremerich J, Flamm SD, Fogel MA, Friedrich MG, et al. Standardized image interpretation and post processing in cardiovascular magnetic resonance: Society for cardiovascular magnetic resonance (scmr) board of trustees task force on standardized post processing. J Cardiovasc Magn Reson. 2013;15:35

27. Dejgaard LA, Skjølsvik ET, Lie ØH, Ribe M, Stokke MK, Hegbom F, et al. The mitral annulus disjunction arrhythmic syndrome. J Am Coll Cardiol. 2018;72:1600–1609

28. Kitkungvan D, Yang EY, El Tallawi KC, Nagueh SF, Nabi F, Khan MA, et al. Extracellular volume in primary mitral regurgitation. JACC. Cardiovascular imaging. 2021;14:1146–1160

29. Messroghli DR, Moon JC, Ferreira VM, Grosse-Wortmann L, He T, Kellman P, et al. Clinical recommendations for cardiovascular magnetic resonance mapping of t1, t2, t2* and extracellular volume: A consensus statement by the society for cardiovascular magnetic resonance (scmr) endorsed by the european association for cardiovascular imaging (eacvi). J Cardiovasc Magn Reson. 2017;19:75

30. Basso C, Iliceto S, Thiene G, Perazzolo Marra M. Mitral valve prolapse, ventricular arrhythmias, and sudden death. Circulation. 2019;140:952–964

31. Hourdain J, Clavel MA, Deharo J-C, Asirvatham S, Avierinos JF, Habib G, et al. Common phenotype in patients with mitral valve prolapse who experienced sudden cardiac death. Circulation. 2018;138:1067–1069

32. Constant Dit Beaufils AL, Huttin O, Jobbe-Duval A, Senage T, Filippetti L, Piriou N, et al. Replacement myocardial fibrosis in patients with mitral valve prolapse: Relation to mitral regurgitation, ventricular remodeling, and arrhythmia. Circulation. 2021;143:1763–1774

33. Flett AS, Hayward MP, Ashworth MT, Hansen MS, Taylor AM, Elliott PM, et al. Equilibrium contrast cardiovascular magnetic resonance for the measurement of diffuse myocardial fibrosis: Preliminary validation in humans. Circulation. 2010;122:138–144

34. Miller CA, Naish JH, Bishop P, Coutts G, Clark D, Zhao S, et al. Comprehensive validation of cardiovascular magnetic resonance techniques for the assessment of myocardial extracellular volume. Circulation. Cardiovascular imaging. 2013;6:373–383

35. Guglielmo M, Fusini L, Muscogiuri G, Baessato F, Loffreno A, Cavaliere A, et al. T1 mapping and cardiac magnetic resonance feature tracking in mitral valve prolapse. Eur Radiol. 2021;31:1100–1109

36. Miller MA, Devesa A, Robson PM, Liao SL, Pyzik R, El-Eshmawi A, et al. Arrhythmic mitral valve prolapse with only mild or moderate mitral regurgitation: Characterization of myocardial substrate. JACC Clin Electrophysiol. 2023;9:1709–1716

37. Essayagh B, Sabbag A, Antoine C, Benfari G, Batista R, Yang LT, et al. The mitral annular disjunction of mitral valve prolapse: Presentation and outcome. JACC. Cardiovascular imaging. 2021;14:2073–2087

38. Bains S, Tester DJ, Asirvatham SJ, Noseworthy PA, Ackerman MJ, Giudicessi JR. A novel truncating variant in flnc-encoded filamin c may serve as a proarrhythmic genetic substrate for arrhythmogenic bileaflet mitral valve prolapse syndrome. Mayo Clin Proc. 2019;94:906–913

39. Roselli C, Yu M, Nauffal V, Georges A, Yang Q, Love K, et al. Genome-wide association study reveals novel genetic loci: A new polygenic risk score for mitral valve prolapse. European heart journal. 2022;43:1668–1680

40. Zuppiroli A, Mori F, Favilli S, Barchielli A, Corti G, Montereggi A, et al. Arrhythmias in mitral valve prolapse: Relation to anterior mitral leaflet thickening, clinical variables, and color doppler echocardiographic parameters. Am Heart J. 1994;128:919–927

41. Zeitler EP, Poole JE, Albert CM, Al-Khatib SM, Ali-Ahmed F, Birgersdotter-Green U, et al. Arrhythmias in female patients: Incidence, presentation and management. Circ Res. 2022;130:474–495

42. Tastet L, Kwiecinski J, Pibarot P, Capoulade R, Everett RJ, Newby DE, et al. Sex-related differences in the extent of myocardial fibrosis in patients with aortic valve stenosis. JACC. Cardiovascular imaging. 2020;13:699–711

43. Pedram A, Razandi M, Narayanan R, Levin ER. Estrogen receptor beta signals to inhibition of cardiac fibrosis. Mol Cell Endocrinol. 2016;434:57–68

44. Bao AM, Liu RY, van Someren EJ, Hofman MA, Cao YX, Zhou JN. Diurnal rhythm of free estradiol during the menstrual cycle. Eur J Endocrinol. 2003;148:227–232

45. Groeneveld SA, Kirkels FP, Cramer MJ, Evertz R, Haugaa KH, Postema PG, et al. Prevalence of mitral annulus disjunction and mitral valve prolapse in patients with idiopathic ventricular fibrillation. J Am Heart Assoc. 2022;11:e025364

46. Otto CM, Nishimura RA, Bonow RO, Carabello BA, Erwin JP, 3rd, Gentile F, et al. 2020 acc/aha guideline for the management of patients with valvular heart disease: Executive summary: A report of the american college of cardiology/american heart association joint committee on clinical practice guidelines. J Am Coll Cardiol. 2021;77:450–500

47. Vahanian A, Beyersdorf F, Praz F, Milojevic M, Baldus S, Bauersachs J, et al. 2021 esc/eacts guidelines for the management of valvular heart disease. European heart journal. 2022;43:561–632

